# Case-mix adjustment of HCAHPS scores based on national benchmarks in Japan: a multicenter cross-sectional study

**DOI:** 10.1101/2025.11.04.25339329

**Authors:** Takuya Aoki, Masato Matsushima

**Affiliations:** Division of Clinical Epidemiology, Research Center for Medical Sciences, The Jikei University School of Medicine, Tokyo, Japan

**Author notes:** **Corresponding author:** Takuya Aoki, Division of Clinical Epidemiology, Research Center for Medical Sciences, The Jikei University School of Medicine, 3-25-8 Nishishimbashi, Minato-ku, Tokyo, 105-8461, Japan.

**Keywords:** case-mix adjustment, HCAHPS, healthcare quality, patient-centered care, patient experience

## Abstract

**Objectives:** Despite the increasing adoption of patient experience measurement in Asia, including Japan, limited progress has been made in identifying suitable patient-level adjusters and developing statistical methods for case-mix adjustment. This study aimed to develop and evaluate a case-mix adjustment model for the Japanese version of the Hospital Consumer Assessment of Healthcare Providers and Systems (HCAHPS) using nationwide benchmark survey data.

**Methods:** We conducted a multicenter cross-sectional study using data from 83 hospitals across Japan that participated in the 2023 national patient experience survey. Self-administered questionnaires were distributed to adult inpatients (aged ≥20 years) discharged between September and November 2023. The Japanese version of HCAHPS, comprising six composite measures and two global ratings, was used to calculate top-box scores (percentage of most positive responses). Candidate case-mix adjusters included age, gender, self-rated general health, self-rated mental health, and admission through the emergency department. The explanatory power (EP) of each adjuster was calculated as the product of predictive power and the between-versus within-hospital variance ratio. The impact of adjustment was evaluated by comparing unadjusted and adjusted scores and by assessing hospital rank changes using Kendall’s τ.

**Results:** Of 31,028 distributed questionnaires, 13,602 were returned (response rate 43.8%). Self-rated mental and general health showed the highest EP, whereas gender and emergency admission contributed minimally. The mean absolute adjustment of top-box scores ranged from 1.52 to 3.20 points. Case-mix adjustment changed hospital rankings in 7.4–16.5% of all possible pairwise comparisons (Kendall’s τ = 0.67–0.85).

**Conclusions:** This study provides the first evidence-based case-mix adjustment model for HCAHPS scores in Japan. Applying such an adjustment is crucial to ensure fair benchmarking of PX across hospitals and to support data-driven quality improvement and accountability initiatives within Japan’s evolving healthcare quality framework.

## INTRODUCTION

Healthcare quality is a multidimensional construct encompassing several core attributes, including effectiveness, safety, and patient-centeredness.^1^ Among these, patient-centeredness has gained increasing attention as a defining feature of high-quality care, emphasizing the integration of patients’ preferences, needs, and values into clinical decision-making.^1^ Patient experience (PX) has emerged as a standardized and measurable indicator of patient-centeredness. Unlike patient satisfaction, which reflects subjective expectations, PX evaluates whether specific care processes occurred and how patients perceived them. Accordingly, there has been a shift from assessments centered on patient satisfaction toward those focusing on concrete care experiences.^2^ Previous studies have demonstrated that PX is not only a vital component of healthcare quality but is also associated with technical quality measures such as clinical effectiveness and safety, as well as with patient behaviors and health outcomes.^3,4^

To assess PX among inpatients, the Hospital Consumer Assessment of Healthcare Providers and Systems (HCAHPS) was developed in the United States.^5^ The HCAHPS has since been implemented nationwide as a core instrument for measuring PX and is utilized in pay-for-performance programs, public reporting, and quality improvement initiatives.^6^ It has also been widely adopted internationally as a standardized measure of PX. The Japanese version of HCAHPS has been developed and psychometrically validated, showing good reliability and validity across diverse hospital settings.^7^ It is currently the most widely used instrument for evaluating PX in Japan.

Benchmarking quality indicators, including PX, across healthcare institutions provides essential insights for quality improvement.^8^ However, valid inter-hospital comparisons require appropriate statistical adjustment for variations in patient characteristics, known as case-mix adjustment.^9^ In Western countries, case-mix adjustment is routinely applied to PX measures to ensure fair comparisons across facilities. For instance, the national HCAHPS survey in the United States adjusts for patients’ demographic and clinical characteristics.^10^ Despite the increasing adoption of PX measurement in Asia, including Japan, limited progress has been made in identifying suitable patient-level adjusters and developing statistical methods for case-mix adjustment. Due to differences in healthcare delivery systems and patients’ sociodemographic characteristics between Asia and Western countries, a Japan-specific case-mix adjustment approach is necessary.

Therefore, this multicenter cross-sectional study aimed to develop and evaluate a case-mix adjustment model for the Japanese version of HCAHPS using nationwide benchmark survey data. This study represents an initial step toward establishing a robust and standardized approach to adjusting PX scores for fair and meaningful hospital comparisons in Japan.

## METHODS

### Design, setting, and participants

We used data from a multicenter cross-sectional survey conducted in 83 hospitals across Japan between September and November 2023 by the Nihon Hospital Alliance (NHA), a Japanese group purchasing organization. Since 2014, the NHA has conducted annual PX surveys to evaluate and improve patient-centeredness in hospitals. Since 2021, the validated Japanese version of HCAHPS has been employed as the PX measure. The hospitals participating in the 2023 survey were located nationwide and ranged in size from small facilities with 88 beds to large institutions with 800 beds. Among them, 5 hospitals had fewer than 200 beds, 55 had 200–499 beds, and 23 had 500 or more beds. A self-administered questionnaire was distributed in person by hospital staff to patients aged ≥ 20 years who were discharged from participating hospitals during the survey period. Within each hospital, eligible participants were selected using a continuous sampling method based on discharge date, and the number of questionnaires distributed was determined according to hospital bed capacity. Patients unable to respond because of severe physical or mental disorders, as assessed by hospital staff at the time of questionnaire distribution, were excluded. Completed questionnaires were returned by mail or via the internet.

### Measures

#### HCAHPS

We used the Japanese version of HCAHPS to assess PX. This scale was developed with permission from the Agency for Healthcare Research and Quality (AHRQ) and the Centers for Medicare and Medicaid Services (CMS), and a previous study conducted in Japan showed it to have good reliability and validity.^7^ The Japanese version of HCAHPS is a 19-item tool consisting of six composite measures and two global ratings. The composites are communication with nurses (Q1–Q3), communication with doctors (Q5–Q7), responsiveness of hospital staff (Q4 and 11), hospital environment (Q8 and 9), communication about medicines (Q13 and 14), and discharge information (Q16 and 17). The global ratings include overall hospital rating (Q18) and willingness to recommend the hospital to friends and family (recommended hospital) (Q19).

The HCAHPS survey employs several response formats: a dichotomous scale (1 = Yes, 2 = No), a global rating scale (0 = Worst to 10 = Best), and two four-point Likert scales (1 = Never, 2 = Sometimes, 3 = Usually, 4 = Always; and 1 = Definitely no, 2 = Probably no, 3 = Probably yes, 4 = Definitely yes). For each item, patient responses were converted into top-box scores, representing the percentage of respondents who selected the most positive response option. For frequency-type questions (e.g., “Never,” “Sometimes,” “Usually,” “Always”), the top-box corresponds to “Always.” For yes/no questions, the top-box is defined as the response “Yes.” For global rating questions using a 0–10 scale (Q18), the top-box is defined as ratings of 9 or 10. For the recommendation item (Q19), the top-box is defined as “Definitely yes.” Composite scores were calculated as the mean of the top-box percentages across the items comprising each domain, consistent with the original HCAHPS scoring methodology.^10^ Top-box scores range from 0 to 100, with higher scores indicating better PX.

#### Case-mix adjusters

Potential case-mix adjusters were selected from data collected in the 2023 PX survey based on the guidelines and previous studies examining the adjustment of PX scores.^10–15^ The following variables were included: age, gender, self-rated general health status, self-rated mental health status, and admission through the emergency department. All adjusters were assessed using a self-administered questionnaire.

### Statistical analysis

To identify case-mix adjusters that were important predictors of PX scores and whose distributions varied across hospitals, we first calculated the explanatory power (EP) for each potential adjuster.^13,15^ EP was defined as the product of predictive power and the variance ratio. To estimate predictive power, null linear regression models were first fitted for each HCAHPS score, including hospital fixed effects to account for clustering of patients within hospitals. Each potential case-mix adjuster was then added individually to these models, and the resulting change in R^2^ was taken as an indicator of predictive power. For each adjuster, the ratio of between-hospital to within-hospital variance was also calculated and multiplied by 1,000 for ease of interpretation.

Two primary approaches were used to assess the impact of case-mix adjustment.^13,15^ First, unadjusted and adjusted top-box scores were calculated for each hospital and each measure. Adjusted scores were derived from a linear regression model including case-mix adjusters as covariates and hospital fixed effects, by averaging predicted values for each hospital with all adjusters set to their reference levels. The difference between adjusted and unadjusted scores was then examined, and we report the largest positive adjustment (adjusted > unadjusted), the largest negative adjustment (adjusted < unadjusted), and the mean absolute adjustment across hospitals. Second, the effect of adjustment on hospital rankings was evaluated using Kendall’s τ correlation coefficient and the proportion of all possible hospital pairs whose rank order changed following adjustment, calculated as ([1 - Kendall’s τ]/2) × 100. Kendall’s τ ranges from -1 to 1, where 1 indicates identical rankings before and after adjustment, and -1 indicates a complete reversal. Statistical analyses were conducted using R, version 4.5.1 (R Foundation for Statistical Computing, Vienna, Austria; www.R-project.org). For missing data, we performed a complete case analysis.

## RESULTS

### Summary of participants’ characteristics

A total of 31,028 questionnaires were distributed, and 13,602 were returned and included in the analysis (response rate: 43.8%). Participants’ characteristics are presented in Table 1. The age distribution was skewed toward older adults: 43.4% were aged 70 years or older, whereas only 9.8% were younger than 40 years. Regarding gender, male and female participants were almost equally represented, with 0.9% preferring not to answer. Most participants rated their general and mental health as fair or good, while only a small proportion rated them as excellent. About one-quarter of participants (27.5%) were admitted through the emergency department. The proportion of missing data ranged from approximately 10% to 12% across variables.

**Table 1.** Participants’ characteristics (n = 13,602)

| Characteristic | n (%) |
| --- | --- |
| Age (year) |  |
| 20–29 | 474 (3.5) |
| 30–39 | 851 (6.3) |
| 40–49 | 956 (7.0) |
| 50–59 | 1479 (10.9) |
| 60–69 | 2282 (16.8) |
| 70–79 | 3740 (27.5) |
| 80 or more | 2158 (15.9) |
| Data missing | 1662 (12.2) |
| Gender |  |
| Male | 6086 (44.7) |
| Female | 6000 (44.1) |
| Prefer not to answer | 119 (0.9) |
| Data missing | 1397 (10.3) |
| Self-rated general health status |  |
| Excellent | 430 (3.2) |
| Very good | 1827 (13.4) |
| Good | 3275 (24.1) |
| Fair | 5049 (37.1) |
| Poor | 1382 (10.2) |
| Data missing | 1639 (12.0) |
| Self-rated mental health status |  |
| Excellent | 779 (5.7) |
| Very good | 2316 (17.0) |
| Good | 3488 (25.6) |
| Fair | 4821 (35.4) |
| Poor | 712 (5.2) |
| Data missing | 1486 (10.9) |
| Admission through emergency department |  |
| Yes | 3740 (27.5) |
| No | 8308 (61.1) |
| Data missing | 1554 (11.4) |

### Explanatory power of case-mix adjusters

The mean predictive power, variance ratio, and EP for the case-mix adjusters are presented in Table 2. Self-rated mental health status showed the highest mean EP across the 17 items and two global ratings, followed by self-rated general health status. In contrast, gender had the lowest mean EP for both the items and global ratings.

**Table 2.** Predictive power, variance ratio, and explanatory power of case-mix adjusters.

|  | Mean |  |  |
| --- | --- | --- | --- |
|  | Predictive power<sup>a</sup> | Variance ratio<sup>b</sup> | Explanatory power<sup>c</sup> |
| Items (17) |  |  |  |
| Age | 0.0043 | 103.0 | 0.46 |
| Gender | 0.0005 | 30.7 | 0.01 |
| Self-rated general health status | 0.0189 | 23.2 | 0.48 |
| Self-rated mental health status | 0.0259 | 21.6 | 0.59 |
| Admission through emergency department | 0.0020 | 60.0 | 0.13 |
| Global ratings (2) |  |  |  |
| Age | 0.0006 | 104.0 | 0.06 |
| Gender | 0.0004 | 27.6 | 0.01 |
| Self-rated general health status | 0.0331 | 20.6 | 0.68 |
| Self-rated mental health status | 0.0500 | 18.4 | 0.92 |
| Admission through emergency department | 0.0002 | 55.9 | 0.01 |

<sup>a</sup>Increase in R<sup>2</sup> from adding the adjuster to a model with hospital fixed effects.
<sup>b</sup>Ratio of between-hospital variance to within-hospital variance (multiplied by 1,000 for presentation).
<sup>c</sup>Predictive power × Variance ratio.

### Impact of case-mix adjustment

As presented in Table 3, the mean absolute adjustment of the top-box scores ranged from 1.52 to 3.20. The largest absolute changes were observed in Communication about medicines, Responsiveness of hospital staff, and Communication with nurses. The greatest negative adjustment was -13.50 for Communication about medicines, whereas the greatest positive adjustment was 27.59 for the same measure. The impact of case-mix adjustment on the relative ranking of hospitals is also shown in Table 3. The Kendall’s τ coefficients ranged from 0.67 to 0.85, indicating that adjustment altered hospital rankings in 7.4–16.5% of all possible pairwise comparisons.

**Table 3.** Impact of case-mix adjustment on top-box scores for composites and global ratings.

|  | Mean (SD)<sup>a</sup> | Changes in top-box scores |  |  | Changes in hospital rankings |  |
| --- | --- | --- | --- | --- | --- | --- |
| | | Mean absolute adjustment | Largest positive adjustment | Largest negative adjustment | Kendall $\tau$ correlation | Percent change<sup>b</sup> |
| Composites |  |  |  |  |  |  |
| Communication with nurses | 65.4 (6.7) | 2.26 | 7.37 | -9.65 | 0.72 | 13.9 |
| Communication with doctors | 71.4 (6.7) | 2.19 | 9.71 | -10.05 | 0.67 | 16.5 |
| Responsiveness of hospital staff | 66.0 (8.8) | 2.39 | 8.34 | -8.20 | 0.74 | 12.9 |
| Hospital environment | 53.6 (7.9) | 1.57 | 6.28 | -3.59 | 0.85 | 7.7 |
| Communication about medicines | 58.6 (10.0) | 3.20 | 27.59 | -13.50 | 0.77 | 11.4 |
| Discharge information | 81.5 (6.3) | 1.52 | 16.76 | -4.71 | 0.79 | 10.4 |
| Global ratings |  |  |  |  |  |  |
| Overall hospital rating | 59.7 (9.3) | 1.95 | 8.17 | -10.93 | 0.85 | 7.4 |
| Recommend hospital | 33.1 (9.5) | 1.66 | 5.81 | -6.98 | 0.85 | 7.4 |

<sup>a</sup>Mean (SD) before case-mix adjustment.<sup>b</sup>Percent of hospital pairs that changed ranking after adjustment.

Figure 1 and Supplementary Table 1 present the distributions of adjusted HCAHPS scores, whereas Supplementary Table 2 presents the distributions of unadjusted HCAHPS scores.

**Figure 1.**
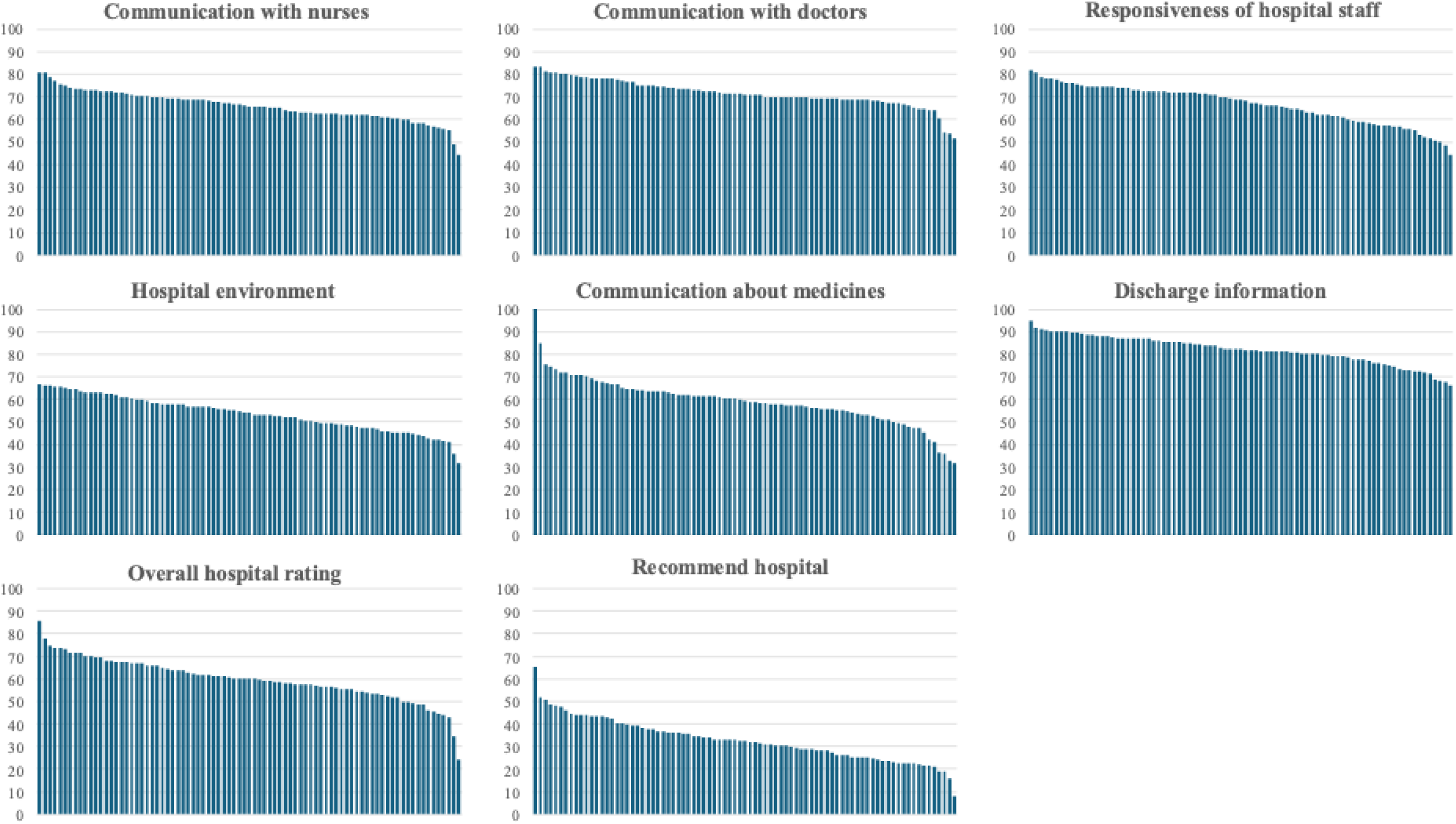
Distribution of case-mix–adjusted HCAHPS top-box scores across 83 hospitals in Japan

## DISCUSSION

### Statement of principal findings

This study represents the first nationwide investigation in Japan to develop and evaluate a case-mix adjustment model for PX measurement. Using data from 83 hospitals across the country, we examined the impact of adjustment on HCAHPS scores and hospital rankings. Our findings demonstrated that self-rated general and mental health status were the strongest case-mix adjusters, whereas gender and admission through the emergency department contributed less EP. Case-mix adjustment altered hospital rankings by 7.4–16.5% of all possible pairwise comparisons, underscoring the importance of incorporating such adjustment to ensure fair benchmarking. This study provides foundational evidence for applying statistically robust case-mix adjustment to PX measurement in Japan, enabling more meaningful hospital comparisons and promoting data-driven quality improvement.

### Strengths and limitations

A major strength of this study is its large-scale, multicenter design encompassing 83 hospitals across Japan, which provides a diverse sample of hospital inpatients. The participating institutions varied in size, region, and type, thereby enhancing the generalizability of the findings. Furthermore, the age and gender distribution of our study participants was consistent with the national trends for hospitalized patients in Japan, suggesting a high degree of representativeness for the entire inpatient population.^16^ Another strength lies in our analytical rigor. We adopted a case-mix adjustment approach based on the statistical methods used in previous international PX measurement studies.

There are several limitations that warrant consideration. First, although the study had a large sample size, the response rate was modest, which could introduce non-response bias. Patients with lower health literacy or poorer experiences may have been less likely to participate. Second, the hospitals that participated in the survey were members of a group purchasing organization that was actively interested in patient-centered quality improvement. Therefore, the findings may not fully represent institutions that are less engaged in quality initiatives. Third, some potential case-mix variables, such as education level, income, primary language, and race/ethnicity, were unavailable in our dataset, which may limit the comprehensiveness of the adjustment.

### Interpretation within the context of the wider literature

Our results are broadly consistent with findings from studies in the United States and other Western countries where the HCAHPS or similar PX instruments have been employed. Prior research has shown that self-rated general and mental health are among the strongest predictors of PX scores, while demographic factors such as age and gender have relatively smaller effects.^12,13,15^ The relatively low EP of age, gender, and admission through the emergency department in our analysis aligns with this evidence. Nonetheless, we retained these variables in our final adjustment model because there is no universally defined threshold of EP that determines inclusion. Moreover, these variables are frequently included in case-mix models for comparability, including the original HCAHPS in the United States.^10^

A notable distinction between our study and previous research is the absence of socioeconomic variables as a case-mix adjuster. For example, education has consistently been used in the original HCAHPS model and in studies using the other PX instruments because it is associated with response tendencies.^9,11,13,14,17^ However, education data were not collected in the NHA survey and were therefore unavailable in this study. This limitation underscores the importance of incorporating socioeconomic variables in future Japanese PX surveys to improve the precision of case-mix adjustment.

### Implications for policy, practice and research

The findings of this study have several implications for the future use of PX data in Japan. As the measurement of PX becomes increasingly integrated into national quality initiatives and policy frameworks,^18^ ensuring equitable hospital comparisons will be essential. Case-mix adjustment offers a statistically grounded means of distinguishing true differences in care quality from variations in patient populations. Our results provide a reference for healthcare administrators, policymakers, and researchers seeking to establish standardized benchmarking systems. In practice, adopting an evidence-based case-mix adjustment model could enhance the credibility and acceptance of PX results among hospitals, thereby encouraging their use in internal audits and quality improvement activities. Furthermore, the dissemination of adjusted PX data may contribute to Japan’s broader efforts toward transparency and accountability in healthcare.

### Conclusions

This study developed and evaluated the first case-mix adjustment model for HCAHPS scores in Japan using data from 83 hospitals nationwide. Our findings highlight the importance of applying case-mix adjustment to ensure fair and accurate PX benchmarking. As Japan moves toward broader PX measurement adoption within its healthcare quality agenda, incorporating robust adjustment methods will be essential for producing reliable indicators that promote quality improvement, accountability, and patient-centered care.

## Contributorship

All authors contributed to the conception or design of the work and to the discussion of the data, reviewed and edited the manuscript, performed critical review of the manuscript, interpreted the analyses and gave the final approval of the manuscript before submission. Dr Aoki performed the statistical analyses and drafted the manuscript. Dr Aoki is the guarantor of the work and accepts full responsibility for the presented content.

## Ethics and other permissions

The institutional review board of the Jikei University School of Medicine provided ethical approval for this study (approval no. 36-387(12505)).

## Funding

This work was supported by JSPS KAKENHI Grant Number JP23K16269.

## Conflict of interests

Drs Aoki and Matsushima received lecture fees and lecture travel fees from the Centre for Family Medicine Development of Japanese Health and Welfare Co-operative Federation. Drs Aoki and Matsushima are advisers of the Centre for Family Medicine Development practice-based research network. Dr. Matsushima’s son-in-law worked at IQVIA Services Japan K.K., which is a contract research organization and a contract sales organization. Dr. Matsushima’s son-in-law works at Syneos Health Clinical K.K., which is a contract research organization and a contract sales organization.

## Acknowledgments

The authors would like to thank Toshio Goto and Yoko Endo (Nihon Hospital Alliance) for their assistance with the administration of this study. The authors disclose the use of ChatGPT for grammar and style checking during the manuscript preparation process. All suggestions were manually reviewed and approved by the authors.

## Data Availability Statement

The data underlying this article were provided by the Nihon Hospital Alliance under license / by permission. Data will be shared on request to the corresponding author with permission of the Nihon Hospital Alliance.

