## Supplementary file for "Case-mix adjustment of HCAHPS scores based on national benchmarks in Japan: a multicenter cross-sectional study"

**Supplementary table 1. Percentile distribution of case-mix–adjusted HCAHPS composite and item-level top-box scores**

| Composites / Measures | Minimum score | 10th percentile | 20th percentile | 30th percentile | 40th percentile | 50th percentile | 60th percentile | 70th percentile | 80th percentile | 90th percentile | Maximum score |
|---|---|---|---|---|---|---|---|---|---|---|---|
| **Communication with nurses** | **44.2** | **58.5** | **61.5** | **62.5** | **63.8** | **65.8** | **68.4** | **69.7** | **71.6** | **73.3** | **80.7** |
| Q1. During this hospital stay, how often did nurses treat you with courtesy and respect? | 51.3 | 61.5 | 63.3 | 65.7 | 67.7 | 68.9 | 71.5 | 73.3 | 74.4 | 76.5 | 85.5 |
| Q2. During this hospital stay, how often did nurses listen carefully to you? | 41.9 | 57.8 | 60.9 | 62.6 | 64.9 | 65.8 | 68.1 | 69.9 | 72.0 | 74.1 | 88.4 |
| Q3. During this hospital stay, how often did nurses explain things in a way you could understand? | 39.6 | 55.6 | 58.7 | 60.5 | 62.0 | 63.5 | 64.1 | 65.9 | 69.0 | 71.8 | 81.0 |
| **Communication with doctors** | **51.8** | **65.2** | **68.5** | **69.3** | **69.8** | **71.0** | **72.7** | **74.6** | **77.4** | **79.1** | **83.5** |
| Q5. During this hospital stay, how often did doctors treat you with courtesy and respect? | 55.4 | 66.7 | 70.7 | 72.8 | 73.7 | 74.9 | 76.1 | 78.5 | 80.8 | 82.4 | 88.4 |
| Q6. During this hospital stay, how often did doctors listen carefully to you? | 48.6 | 65.5 | 66.9 | 67.9 | 69.2 | 70.5 | 71.7 | 74.0 | 76.5 | 78.7 | 82.8 |
| Q7. During this hospital stay, how often did doctors explain things in a way you could understand? | 44.0 | 62.6 | 64.4 | 66.8 | 68.0 | 69.3 | 70.6 | 72.9 | 75.2 | 77.2 | 81.2 |
| **Responsiveness of hospital staff** | **44.2** | **55.8** | **58.8** | **62.0** | **65.7** | **68.9** | **71.5** | **72.3** | **74.3** | **75.9** | **81.7** |
| Q4. During this hospital stay, after you pressed the call button, how often did you get help as soon as you wanted it? | 43.0 | 55.2 | 58.5 | 62.1 | 67.1 | 69.2 | 72.0 | 73.5 | 75.5 | 77.4 | 83.7 |
| Q11. How often did you get help in getting to the bathroom or in using a bedpan as soon as you wanted? | 15.9 | 45.5 | 54.7 | 58.7 | 60.9 | 63.1 | 64.8 | 67.2 | 70.1 | 74.6 | 82.0 |
| **Hospital environment** | **31.6** | **44.4** | **47.1** | **49.3** | **52.2** | **54.3** | **56.6** | **58.1** | **60.9** | **63.3** | **66.7** |
| Q8. During this hospital stay, how often were your room and bathroom kept clean? | 44.7 | 50.5 | 58.3 | 61.5 | 64.1 | 66.4 | 69.2 | 70.9 | 75.8 | 78.3 | 84.4 |
| Q9. During this hospital stay, how often was the area around your room quiet at night? | 18.8 | 30.9 | 34.1 | 36.9 | 39.5 | 41.1 | 43.9 | 45.6 | 48.9 | 53.4 | 60.2 |
| **Communication about medicines** | **31.5** | **47.6** | **52.8** | **55.8** | **57.5** | **59.2** | **61.6** | **63.4** | **65.9** | **70.8** | **103.2** |
| Q13. Before giving you any new medicine, how often did hospital staff tell you what the medicine was for? | 41.6 | 58.4 | 63.0 | 66.7 | 68.3 | 70.4 | 72.7 | 75.6 | 78.1 | 82.1 | 104.3 |
| Q14. Before giving you any new medicine, how often did hospital staff describe possible side effects in a way you could understand? | 14.8 | 34.9 | 40.2 | 44.1 | 46.2 | 48.0 | 49.6 | 51.5 | 56.4 | 59.6 | 102.3 |
| **Discharge information** | **65.9** | **72.7** | **77.3** | **79.9** | **81.2** | **82.1** | **84.3** | **85.8** | **87.6** | **89.9** | **95.0** |
| Q16. During this hospital stay, did doctors, nurses or other hospital staff talk with you about whether you would have the help you needed when you left the hospital? | 71.5 | 78.8 | 82.4 | 84.3 | 86.0 | 86.3 | 86.9 | 88.2 | 89.0 | 91.7 | 96.7 |
| Q17. During this hospital stay, did you get information in writing about what symptoms or health problems to look out for after you left the hospital? | 50.1 | 66.1 | 70.1 | 73.2 | 76.0 | 78.7 | 82.1 | 85.3 | 87.1 | 90.3 | 95.6 |
| **Overall hospital rating** | **24.4** | **48.9** | **53.5** | **56.2** | **58.0** | **60.1** | **61.5** | **64.6** | **67.4** | **71.1** | **85.8** |
| **Recommend hospital** | **7.8** | **22.4** | **24.6** | **27.7** | **30.1** | **32.4** | **34.1** | **36.8** | **40.4** | **44.0** | **65.1** |

**Supplementary table 2. Percentile distribution of unadjusted HCAHPS composite and item-level top-box scores**

| Composites / Measures | Minimum score | 10th percentile | 20th percentile | 30th percentile | 40th percentile | 50th percentile | 60th percentile | 70th percentile | 80th percentile | 90th percentile | Maximum score |
|---|---|---|---|---|---|---|---|---|---|---|---|
| **Communication with nurses** | **38.6** | **58.4** | **61.0** | **62.4** | **63.7** | **65.9** | **67.3** | **69.7** | **71.3** | **72.9** | **77.4** |
| Q1. During this hospital stay, how often did nurses treat you with courtesy and respect? | 43.1 | 60.7 | 63.2 | 64.8 | 66.9 | 69.6 | 71.2 | 72.7 | 74.5 | 76.1 | 82.2 |
| Q2. During this hospital stay, how often did nurses listen carefully to you? | 37.3 | 56.2 | 59.9 | 61.6 | 63.9 | 66.1 | 68.1 | 69.5 | 72.3 | 74.1 | 78.1 |
| Q3. During this hospital stay, how often did nurses explain things in a way you could understand? | 35.3 | 55.0 | 58.2 | 59.4 | 61.1 | 63.1 | 65.2 | 66.4 | 68.3 | 70.9 | 74.1 |
| **Communication with doctors** | **41.8** | **64.3** | **67.4** | **69.2** | **69.9** | **71.6** | **73.3** | **75.4** | **77.3** | **78.9** | **83.1** |
| Q5. During this hospital stay, how often did doctors treat you with courtesy and respect? | 46.0 | 66.5 | 71.0 | 72.0 | 73.6 | 74.5 | 76.6 | 78.2 | 80.3 | 82.6 | 90.1 |
| Q6. During this hospital stay, how often did doctors listen carefully to you? | 40.0 | 63.3 | 66.1 | 67.6 | 69.6 | 70.8 | 72.7 | 74.2 | 76.9 | 78.3 | 81.5 |
| Q7. During this hospital stay, how often did doctors explain things in a way you could understand? | 40.0 | 61.3 | 64.5 | 66.7 | 68.3 | 69.6 | 70.8 | 73.5 | 75.5 | 77.2 | 81.4 |
| **Responsiveness of hospital staff** | **41.2** | **54.5** | **57.4** | **61.8** | **65.0** | **67.9** | **69.7** | **71.8** | **72.8** | **75.7** | **82.5** |
| Q4. During this hospital stay, after you pressed the call button, how often did you get help as soon as you wanted it? | 40.6 | 54.5 | 58.4 | 61.6 | 65.7 | 68.6 | 70.2 | 73.1 | 74.1 | 78.4 | 85.9 |
| Q11. How often did you get help in getting to the bathroom or in using a bedpan as soon as you wanted? | 16.7 | 43.2 | 49.3 | 55.1 | 60.0 | 62.8 | 64.9 | 66.7 | 69.2 | 71.9 | 90.0 |
| **Hospital environment** | **33.7** | **43.8** | **46.7** | **48.9** | **51.6** | **53.8** | **56.5** | **58.0** | **61.1** | **63.6** | **69.0** |
| Q8. During this hospital stay, how often were your room and bathroom kept clean? | 45.0 | 50.1 | 57.6 | 61.5 | 63.5 | 66.1 | 67.9 | 70.5 | 74.9 | 77.7 | 85.0 |
| Q9. During this hospital stay, how often was the area around your room quiet at night? | 21.8 | 30.6 | 34.0 | 36.6 | 39.0 | 41.2 | 43.9 | 46.2 | 50.0 | 53.9 | 58.3 |
| **Communication about medicines** | **31.3** | **46.1** | **50.8** | **54.6** | **56.9** | **59.9** | **60.7** | **63.3** | **67.0** | **70.9** | **77.4** |
| Q13. Before giving you any new medicine, how often did hospital staff tell you what the medicine was for? | 46.2 | 54.1 | 59.9 | 65.2 | 67.2 | 70.6 | 72.6 | 76.4 | 77.7 | 82.3 | 89.3 |
| Q14. Before giving you any new medicine, how often did hospital staff describe possible side effects in a way you could understand? | 12.5 | 36.5 | 40.0 | 42.1 | 46.5 | 48.3 | 49.7 | 51.3 | 54.5 | 60.5 | 70.0 |
| **Discharge information** | **63.2** | **72.1** | **75.8** | **79.0** | **80.8** | **82.2** | **83.7** | **85.2** | **87.3** | **89.0** | **92.6** |
| Q16. During this hospital stay, did doctors, nurses or other hospital staff talk with you about whether you would have the help you needed when you left the hospital? | 71.7 | 78.9 | 81.1 | 83.5 | 85.0 | 86.3 | 87.3 | 88.0 | 88.7 | 91.0 | 97.9 |
| Q17. During this hospital stay, did you get information in writing about what symptoms or health problems to look out for after you left the hospital? | 44.2 | 65.7 | 69.9 | 71.7 | 74.7 | 78.1 | 81.2 | 85.0 | 86.5 | 90.0 | 94.2 |
| **Overall hospital rating** | **35.3** | **49.0** | **54.4** | **55.6** | **57.9** | **59.5** | **62.3** | **65.7** | **68.5** | **72.1** | **77.6** |
| **Recommend hospital** | **6.3** | **22.0** | **24.1** | **27.1** | **30.5** | **32.9** | **34.9** | **37.9** | **41.7** | **44.6** | **64.9** |
